# Consensus Guideline for the Management of Gastric Cancer with Synchronous Peritoneal Metastases

**DOI:** 10.1101/2024.04.10.24305456

**Authors:** PSM Writing Group, PSM Consortium Group, Kiran K Turaga

**Affiliations:** Yale New Haven Health

## Abstract

**Background:** Gastric cancer with synchronous peritoneal metastases (GCPM) is a debilitating disease with limited treatment options. This manuscript describes an update of the 2018 Chicago Consensus Guidelines addressing the management of GCPM in line with most recent evidence.

**Methods:** A clinical management pathway was updated through two rounds of a Delphi Consensus to assess agreement levels with pathway blocks. Supporting evidence underwent evaluation via a rapid literature review. Meta-analyses were performed where appropriate.

**Results:** Overall, level of evidence in this disease subset was low to moderate. Of 124 participants in the first round, 109 (88%) responded in the second round. Strong consensus (>90%) was achieved in 6/8 (75%) blocks in round I and II. A multidisciplinary preoperative assessment and diagnostic laparoscopy should be offered all patients, while patients with a high burden of disease or progression should undergo non-surgical management. Patients with stable/responsive disease and low peritoneal carcinomatosis index should subsequently be offered treatment with regional therapeutic interventions and cytoreductive surgery. In patients who are cytology positive, systemic therapy can be used to convert these patients to cytology negative, with subsequent surgery offered per the patient’s goals of care. Meta-analysis of observational and randomized control trials revealed a survival benefit with the addition of intraperitoneal chemotherapy to cytoreductive surgery (HR 0.52).

**Conclusion:** The consensus-driven clinical pathway for GCPMs offers vital clinical guidance for practitioners. There is a growing body of high-quality evidence to support management strategies and future clinical trials are eagerly awaited.

## INTRODUCTION

Gastric cancer is the fifth leading cause of cancer mortality worldwide and was responsible for just under one million new cases in 2022, ranking fifth for incidence.^1^ In the US, up to 65% of patients present with stage III or IV disease, and in those with metastatic disease, the peritoneum is a common site of metastatic spread.^2,3^ Traditionally, stage IV gastric cancer is not a surgical disease. While advancements in systemic and regional therapeutic interventions hold promise, there are several matters of equipoise and variations between institutional practices regarding gastric cancer with peritoneal metastases (GCPM).^4^

Considering this lack of standardization, consensus guidelines on the management of GCPM were created in 2018 as part of the Chicago Consensus Working Group.^5^ Since the inception of these guidelines, there have been major advancements in systemic and regional interventions for GCPM and cytology positive gastric cancer. Herein, we present updated recommendations including a revised clinical management pathway supported by evidence from rapid systematic reviews.

## METHODS

This initiative was part of a national multidisciplinary consortium group process aimed at streamlining guidelines for the care of patients with peritoneal surface malignancies (PSM). The consensus and rapid review methodology has been described in detail in a separate manuscript [submitted].^6^ Major components are summarized below. Search strategies for the scoping review can be seen in Figure 4.

### Consensus Group Structure

In brief, the Gastric Cancer Working Group consisting of seven experts appointed to lead the section, and each pathway iteration was reviewed by the Steering Committee. Two core group members (SDB, VB) coordinated the effort. A team of nine surgical residents and surgical oncology fellows conducted the rapid reviews.

### Modified Delphi Process

The original Chicago Consensus guidelines were first reviewed and revised by the Gastric Cancer Working Group and the Consortium leadership to align with evidence published since the last consensus. Recommendations were revised using a Delphi method across the entire PSM Consortium group by soliciting degrees of agreement with each recommendation on a five-point Likert scale via Qualtrics survey. A threshold of 75% was set to retain a given guideline subject, and 90% to finalize a guideline.

Two Delphi rounds were conducted; at the conclusion of each, the results of the prior round were collected and analyzed, and revisions proposed by the disease site working groups. Voting eligibility was first screened by participation in both Delphi rounds; only those who voted in Delphi 1 qualified to vote in Delphi 2. Levels of evidence were assigned to pathway blocks. Simultaneously, a summary table outlining first-line systemic therapies for GCPM was generated in conjunction with medical oncologists in the Working Group.

### Rapid Review of Literature

A MEDLINE search via PubMed between January 2000 and August 2023 addressed the following key questions

1. KQ 1: What is the optimal management strategy for peritoneal cytology positive gastric cancer without clinically evident peritoneal carcinomatosis?^7^
2. KQ 2: What regional (intraperitoneal) therapeutic interventions are effective in the management of GCPM?^8^

Search strategies were peer-reviewed by a medical librarian specialist and review were registered in PROSPERO before data extraction (CRD42023466035 & CRD42023466032). The Covidence platform facilitated title and abstract screening, full-text review, data extraction, and quality assessment using the Cochrane Risk of Bias 2.0 tool for RCTs and the Newcastle Ottawa Scale for non-randomized studies, respectively.^9–12^ References from relevant articles were searched and reviewed manually by two reviewers. The review was conducted in alignment with recommendations from the Cochrane Rapid Review Methods Groups and reported in line with the Preferred Reporting Items for Systematic Reviews and Meta-Analyses (PRIMSA) 2020 guidelines.^13^

### External Perspectives

Patients advocates within the Hope for Stomach Cancer (STOCAN) organization reviewed the treatment pathway and offered insights regarding clinical trial enrollment, research outcomes, and available resources for patients with GCPMs. Additionally, members of the Peritoneal Surface Oncology Group International (PSOGI) Executive Council were invited to appraise the second version of the pathway. Their comments were consolidated to evaluate alignment with global practices regarding the management of GCPM.

### Systemic therapy recommendations

A section on systemic therapies for GCPM was included under Block 8 and summarized as a table. This was drafted collaboratively with the working group with particular assistance from the medical oncologists in the group.

## RESULTS

### Pathways

Two pathways were initially proposed, one for synchronous, and the other for metachronous GCPM. However, the latter was not established owing to a lack of evidence. In all, 124 experts and thought leaders voted on the clinical pathway for synchronous GCPM, of which 109 (88%) responded in the second Delphi round. The group included 93 (75%) surgical oncologists, 16 (13%) medical oncologists, 11 (9%) pathologists, and 4 (3%) experts from other specialties. Given the low-moderate quality of existing evidence, many recommendations were based on expert opinion. This pathway was divided into eight main blocks (Figure 1). The results of two rounds of modified Delphi processes are summarized in Tables 2 and 3. Overall, strong consensus (>90%) was achieved in 6/8 (75%) blocks in round I and II.

**Figure 1:**
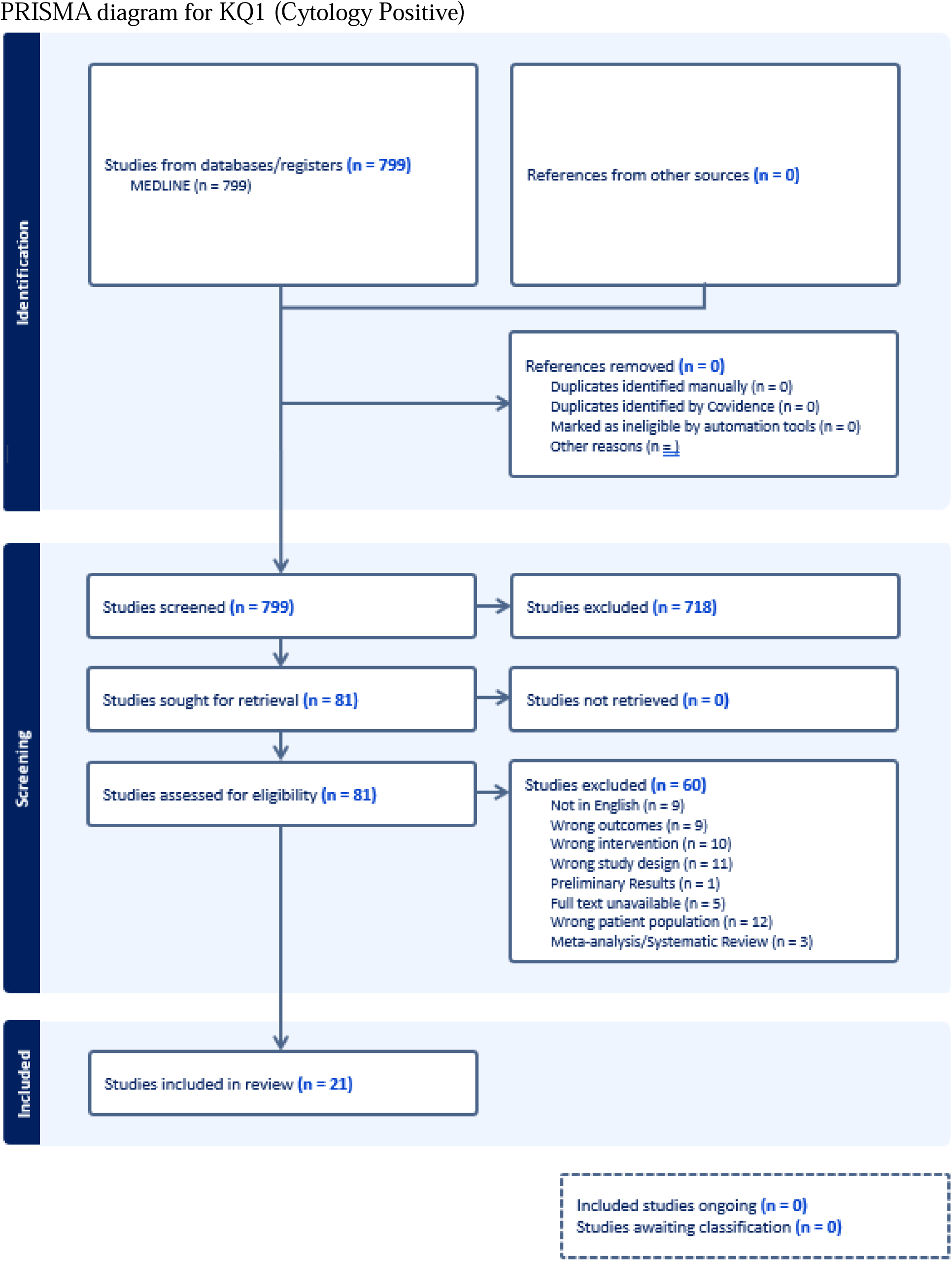
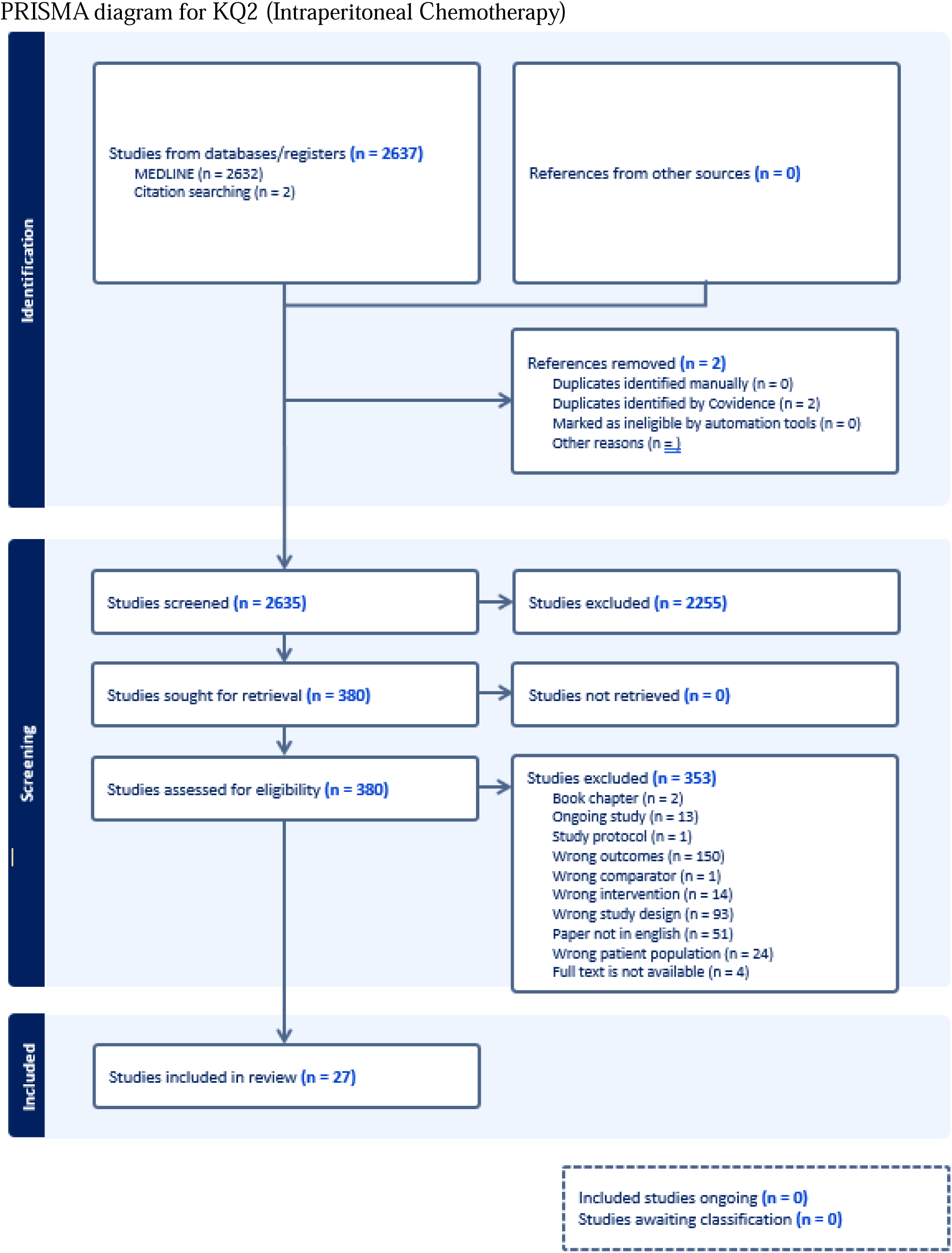
Gastric Cancer with Synchronous Peritoneal Metastasis Clinical Pathway

**Table 1:** Delphi Round 1 agreement table.

|  | **Strongly agree** | **Agree** | **Neither agree nor disagree** | **Disagree** | **Strongly disagree** | **Total** | **Percentage** |
| --- | --- | --- | --- | --- | --- | --- | --- |
| **Block 1** | 90 | 32 | 1 | 1 | 0 | 124 | 98% |
| **Block 2** | 86 | 34 | 3 | 1 | 0 | 124 | 97% |
| **Block 3** | 84 | 36 | 4 | 0 | 0 | 124 | 97% |
| **Block 4** | 89 | 30 | 4 | 1 | 0 | 124 | 96% |
| **Block 5** | 64 | 46 | 9 | 4 | 1 | 124 | 89% |
| **Block 6** | 59 | 44 | 14 | 6 | 1 | 124 | 83% |
| **Block 7** | 79 | 34 | 7 | 3 | 1 | 124 | 91% |
| **Block 8** | 78 | 36 | 7 | 3 | 0 | 124 | 92% |

**Table 2:** Delphi Round 2 agreement table.

|  | **Strongly agree** | **Agree** | **Neither agree nor disagree** | **Disagree** | **Strongly disagree** | **Total** | **Percentage** |
| --- | --- | --- | --- | --- | --- | --- | --- |
| **Block 1** | 99 | 8 | 0 | 2 | 0 | 109 | 98% |
| **Block 2** | 96 | 9 | 3 | 1 | 0 | 109 | 96% |
| **Block 3** | 99 | 7 | 3 | 0 | 0 | 109 | 97% |
| **Block 4** | 103 | 5 | 1 | 0 | 0 | 109 | 99% |
| **Block 5** | 89 | 7 | 9 | 2 | 2 | 109 | 88% |
| **Block 6** | 76 | 17 | 12 | 2 | 2 | 109 | 85% |
| **Block 7** | 94 | 11 | 2 | 2 | 0 | 109 | 96% |
| **Block 8** | 101 | 7 | 1 | 0 | 0 | 109 | 99% |

**Table 3:** Systemic therapies for gastric cancer with peritoneal metastases.

| Performance Status | Therapy | Regimen |
| --- | --- | --- |
| **Karnofsky Performance Score ≥ 60% or ECOG ≤ 2** | *Systemic Therapy<sup>1</sup>* | <u>HER 2 positive</u><br>Fluoropyrimidine, oxaliplatin and trastuzumab +/- pembrolizumab<br><u>HER 2 negative</u><br>Fluoropyrimidine, oxaliplatin, +/- nivolumab/pembrolizumab (PD-L1 CPS ≥ 5)<br><u>MSI-H/dMMR (independent of PD-L1 status)</u><br>Fluoropyrimidine, oxaliplatin, nivolumab/pembrolizumab<br><u>Claudin positive</u><br>Fluoropyrimidine, oxaliplatin and Zolbetuximab (pending FDA approval) |
|  | *Best Supportive Care<sup>2</sup>* | Intended for patients who choose not to receive chemotherapy or for whom the risks of chemotherapy outweigh the benefits. |
| **Karnofsky Performance Score $<$ 60% or ECOG ≥ 3** | *Best Supportive Care* |  |

<sup>1</sup> Universal testing for microsatellite instability (MSI) by PCR/next-generation sequencing (NGS) or mismatch repair (MMR) by immunohistochemistry (IHC) along with HER2 and PD-L1 testing, should be complete if metastatic disease is documented/suspected.
<sup>2</sup> Measures should be taken to support the best possible quality of life for patients and their families. These measures include control of bleeding via interventional radiology, endoscopic treatment, external beam radiation therapy (EBRT) and gastrectomy in select patients; alleviation or bypass of obstructions; pain control; nausea and vomiting control.

### Rapid Review

The first key question regarding cytology positive gastric cancer revealed 799 abstracts for screening. Of these, 81 were considered for full text review and 21 for data extraction. For the second key question about GCPM, we screened 2637 abstracts, of which, 380 were considered for full text review and 27 for data extraction. Relevant exclusion criteria are detailed in the PRISMA flow diagrams. Meta-analysis was performed wherever feasible.

### Summary of Major Changes

The current guidelines, building upon the 2018 Chicago Consensus, feature a more rigorous methodology involving a wider range of experts and patient advocates. They emphasize a thorough preoperative assessment encompassing genetic profiling, psychosocial support, nutrition, fertility considerations, and collaboration with patient advocacy groups. In contrast to the previous guidelines, which recommended direct initiation of standard chemotherapy for six months before restaging, the current pathway advocates for a diagnostic laparoscopy to evaluate the peritoneal cancer index (PCI). For patients with PCI >7-10, systemic therapy, clinical trials, or supportive care are recommended, whereas those with low PCI or positive cytology are advised to undergo systemic therapy with an intent for restaging. After restaging, the pathways converge, with patients who progress receiving additional systemic therapy or supportive care based on functional status and goals. Regional interventions are recommended for patients with stable/responsive disease, including intraperitoneal port-based therapies and cytoreductive surgery (CRS) in cases where complete cytoreduction (CC0) is anticipated. The latter is elaborate upon further with a meta-analysis comparing CRS + intraperitoneal chemotherapy (IPCT) with CRS alone (CRSa).

#### Block 1 (Agreement: Round 1 98%; Round 2 98%)

Preoperative evaluation entails a thorough history and physical exam, including an exploration of the patient’s social history, financial environment, and support networks. Following upper endoscopy and subsequent staging, a CT scan of the abdomen and pelvis with IV contrast should be performed to identify the extent of peritoneal disease and tumor burden.^14^ 18-fluorodeoxyglucose (FDG)-PET is reserved for patients with equivocal findings on CT imaging or patients with clinical indications of metastatic disease and otherwise negative imaging.

High-risk features in advanced gastric cancer, such as tumor size, depth of serosal invasion, perforation, involvement of multiple anatomic regions, and lymph node positivity, warrant close attention, as occult peritoneal metastases may be present in over one-third of patients.^15^ Additionally, poor prognostic indicators identified on imaging like extensive lymph node metastases and obstructive lesions in the biliary, urinary, or gastrointestinal tracts may necessitate alternative management strategies as outlined in block 8.^16^

Any pathology specimens obtained should be tested for EBV, HER2, MSI/MMR status, and PD-L1.^17–20^ Establishing a comprehensive patient support network is highly encouraged and includes patient support, counseling, social work referrals, and early palliative care as indicated. Formal evaluation by a multidisciplinary team or Tumor Board is critical to guide appropriate steps in management.

#### Block 2 (Agreement: Round 1 97%; Round 2 96%)

**A diagnostic laparoscopy is recommended to determine the peritoneal carcinomatosis index (PCI) if preliminary workup reveals low radiographic burden of disease.**^21^ Cytological examination of peritoneal lavage fluid is a key prognostic factor in the classification of gastric carcinoma. Positive cytology is a poor prognostic factor.22 In patients with any M1 disease or positive cytology, NCCN guidelines recommend palliative management. **However, our consensus and pathway recommend proceeding to systemic therapy.**

##### Cytology Positive patients

In patients who are cytology positive with low PCI (< 7), management remains controversial, as positive cytology remains a poor prognostic indicator. Several groups have shown that surgery plus IPC has therapeutic benefit for cytology positive patients, compared to standard therapy or surgery alone (supplemental Tables 3 and 4).^23–36^ In addition, converting patients from positive to negative cytology greatly improves their survival. Given this evidence, initiation of systemic therapy with an intent of restaging is recommended as the first step for patients with positive peritoneal cytology and/or low PCI.

##### PCI Cutoff

Further disagreement exists about the optimal cutoff for low versus high PCI, which are often institution dependent. A systematic review and meta-analysis published in 2015 reported that the median survival changes significantly above a PCI of 12.^37^ The strongest trial representing these results is by Glehen et al., which showed that the best results in terms of survival are patients with a PCI ≤6.^38^

More recently, the CYTO-CHIP study further demonstrated that completeness of CRS was closely linked to tumor burden (PCI).^39^ They showed that long term survival was rare in patients with a PCI > 13. The mean PCI was 7.2 in the CRS-HIPEC group and 2.11 in the CRSa group. **Ultimately, a cutoff ranging from 7-10 was recommended for differentiating low vs. high PCI to determine subsequent selection for regional therapeutic interventions.** However, the anatomic distribution of peritoneal metastasis and histology should be incorporated into the decision-making process. For example, a patient with PCI of 10 and signet ring cell gastric cancer should be advised against cytoreductive surgery given the poor prognosis associated with this histologic type.^40^

#### Block 3 (Agreement: Round 1 97%; Round 2 97%)

Once systemic therapy has concluded, restaging should be performed via computed tomography and diagnostic laparoscopy with peritoneal washings and/or biopsies. Laparoscopy is the gold standard for determining disease response to therapy.

#### Block 4 (Agreement: Round 1 96%; Round 2 99%)

In the presence of disease progression intraperitoneally and/or extra-peritoneally or poor functional status, regional therapeutic interventions are not recommended. Instead, further lines of systemic therapy, enrollment in a clinical trial or supportive care should be initiated. Supportive care can include feeding access or relief obstruction via surgical or endoscopic interventions; control of bleeding; anti-nausea medications; pain relief; initiation of palliative care if not already engaged; hospice resources, etc.

#### Block 5 (Agreement: Round 1 89%; Round 2 88%)

##### Recommendation

**In patients with PCI < 7-10 and disease stable/responsive to chemo, regional therapeutic interventions are recommended. These may include cytoreductive surgery with gastrectomy and D2 lymph node dissection, port based IPCT combined with systemic chemotherapy, and laparoscopic IPCT.**

##### Principles of Surgery

The principles of surgery for patients with GCPM have largely been unchanged since the 2018 guidelines were published. CC0 cytoreduction remains the gold standard and an independent predictor for overall survival in patients undergoing CRS for GCPM.^37^ The extent of gastrectomy is dependent on tumor location and distribution; it has not been shown to be an independent predictor of survival.^41,42^ In patients with locally advanced gastric cancer without peritoneal metastasis, a curative gastrectomy with D2 lymphadenectomy is the standard of care.^43^

CRS has emerged as an important treatment modality for GCPM patients, as systemic therapies have limited effects on peritoneal carcinomatosis likely due to the blood–peritoneal barrier. The theory behind the efficacy of CRS is that debulking allows tumor cells to re-enter the proliferative phase of the cell cycle, potentially becoming more sensitive to anti-neoplastic agents. The goal of cytoreductive surgery is to remove all macroscopic disease, thus achieving “complete cytoreduction.” A CC-0 score indicates that no visible peritoneal seeding exists following cytoreduction. Patient selection remains crucial for CRS, as the extent of disease as measured by PCI can negate the benefit of surgery and IPC. As mentioned in Block 2, a PCI cutoff for surgery of 7-10 should be employed.

##### CRS + Chemotherapy versus Chemotherapy Alone

Several groups have evaluated whether CRS is superior to chemotherapy alone, but only two randomized controlled trials exist. The REGATTA trial examined whether CRS in combination with chemotherapy was superior to chemotherapy alone.^44^ Conducted across three countries, the results failed to demonstrate an overall survival (OS) benefit for the surgical arm. This suggests that incomplete cytoreduction with residual metastatic disease may not confer a survival advantage. Some limitations include a failure to accrue patients and an unbalanced primary tumor location between groups. Given that this trial included patients with extraperitoneal metastasis, it was not included in our rapid review.

The next trial was the GYMSSA trial.^45^ Conducted in the United States, it patients were randomized to systemic chemotherapy or gastrectomy, CRS, HIPEC and systemic chemotherapy (GYMS Arm). The trial demonstrated an improved median OS with complete CRS-HIPEC compared to systemic chemotherapy alone (11.3 months vs. 4.3 months). However, this trial failed to accrue the targeted sample size of 136 patients, precluding robust conclusions.

Five observational studies were included in the rapid review (see Supplemental Table 6).^46–50^ In 2016, Boerner et al. evaluated 38 consecutive GCPM patients that were treated with gastrectomy, CRS, and HIPEC and compared them to 27 patients who received chemotherapy with gastrectomy (PC-Standard).^47^ They found that the CRS-HIPEC group had better overall, 1-year, 3-year and 5-year survival compared to the PC-Standard group. In 2021, Canbay et al. evaluated 53 patients with cytology positive or peritoneal metastases.^46^ All patients underwent laparoscopic HIPEC followed by neoadjuvant intraperitoneal and systemic therapy. 34 of these patients went on to receive CRS and HIPEC, while 19 only underwent induction chemotherapy. The group that underwent CRS-HIPEC had improved overall survival as compared to the chemotherapy alone group (21.2 vs. 15.9 months). Most recently, AkturkEsen et al. showed that patients undergoing CRS-HIPEC after neoadjuvant chemotherapy had improved overall survival as compared to chemotherapy only (19.7 vs. 6.8 months).^50^

##### CRS + IPC versus CRS alone (CRSa)

In addition to evaluating whether the addition of CRS to chemotherapy improves outcomes in GCPM patients, other groups have examined the effect of intraperitoneal chemotherapy alongside CRS. Two randomized controlled trials exist in this space.

The first RCT was published in 2011 by Yang et al.^41^ In this study, 68 patients with GCPM were randomized into CRS alone (n = 34) or CRS-HIPEC (n = 34). Median survival was improved in the CRS-HIPEC group (11.0 vs. 6.5 months), a nearly 70% extension of overall survival. It is worth noting that the median PCI for both groups was 15, which is above our recommended cutoff of 7-10. In addition, this study included metachronous GCPM.

While Yang et al. showed an improved survival with CRS-HIPEC, the GASTRIPEC 1 trial showed no overall survival between CRS-HIPEC and CRS alone.^51^ In this study, patients were randomized to perioperative chemotherapy and CRS alone or CRS-HIPEC. Median survival was the same for both groups (14.9 months). Progression free and metastasis-free survival were significantly better in the CRS-HIPEC group, however. This study ended prematurely because of slow recruitment, and in 55 patients, treatment stopped before CRS mainly due to disease progression and/or death. Importantly, 44% of patients in this study had a PCI ≥ 7 and 40% had ascites, both known factors for poor prognosis after CRS.

Aside from the two RCTs, there have been four observational studies evaluating CRS-HIPEC vs. CRSa (see Supplemental Table 5). The CYTO-CHIP study was a propensity score analysis of patients with GCPM who underwent CRS-HIPEC or CRSa. They showed that CRS-HIPEC had improved overall (18.8 months vs. 12.1) and recurrence-free survival (5.87% vs. 3.76%) as compared to CRSa.^39^ Rosa et al. found that CRS-HIPEC performed for a cure or prophylaxis had better 5-year disease free survival as compared to CRSa. In 2013, Wu et al. found that CRS-HIPEC had improved overall survival as compared to CRSa (15.5 vs. 10.4 months);^52^ of note, they specifically looked at GCPM patients with ovarian metastasis. In 2022, Morgagni et al. found that in patients who received neoadjuvant chemotherapy, CRS-HIPEC patients had improved overall survival as compared to CRSa (46.7 vs. 14.4 months).^53^

##### Meta-Analysis

We performed a meta-analysis comparing CRS-HIPEC to CRSa. Figure 2 shows the results of a meta-analysis evaluating hazard ratios, while Figure 3 shows median overall survival. When evaluating the HR, in both observational studies and randomized controlled there was a HR that favored the addition of HIPEC (HR 0.52 for both). With regard to median overall survival, neither the randomized trials nor the observational studies showed a statistically significant improvement in median overall survival, though several individual trials did.

**Figure 2:**
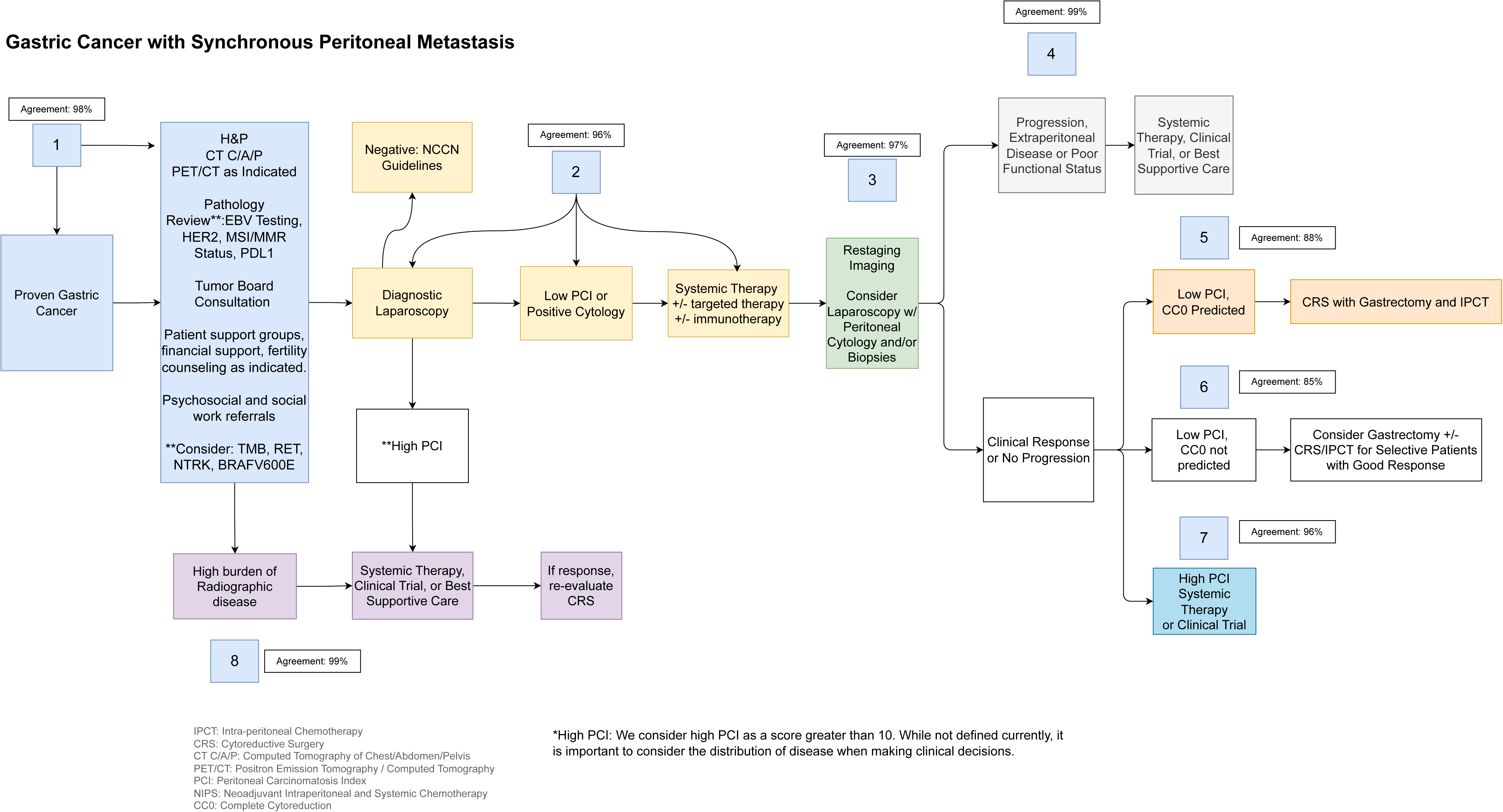
Meta-analysis for hazard ratios comparing CRS-HIPEC to CRS alone

**Figure 3:**
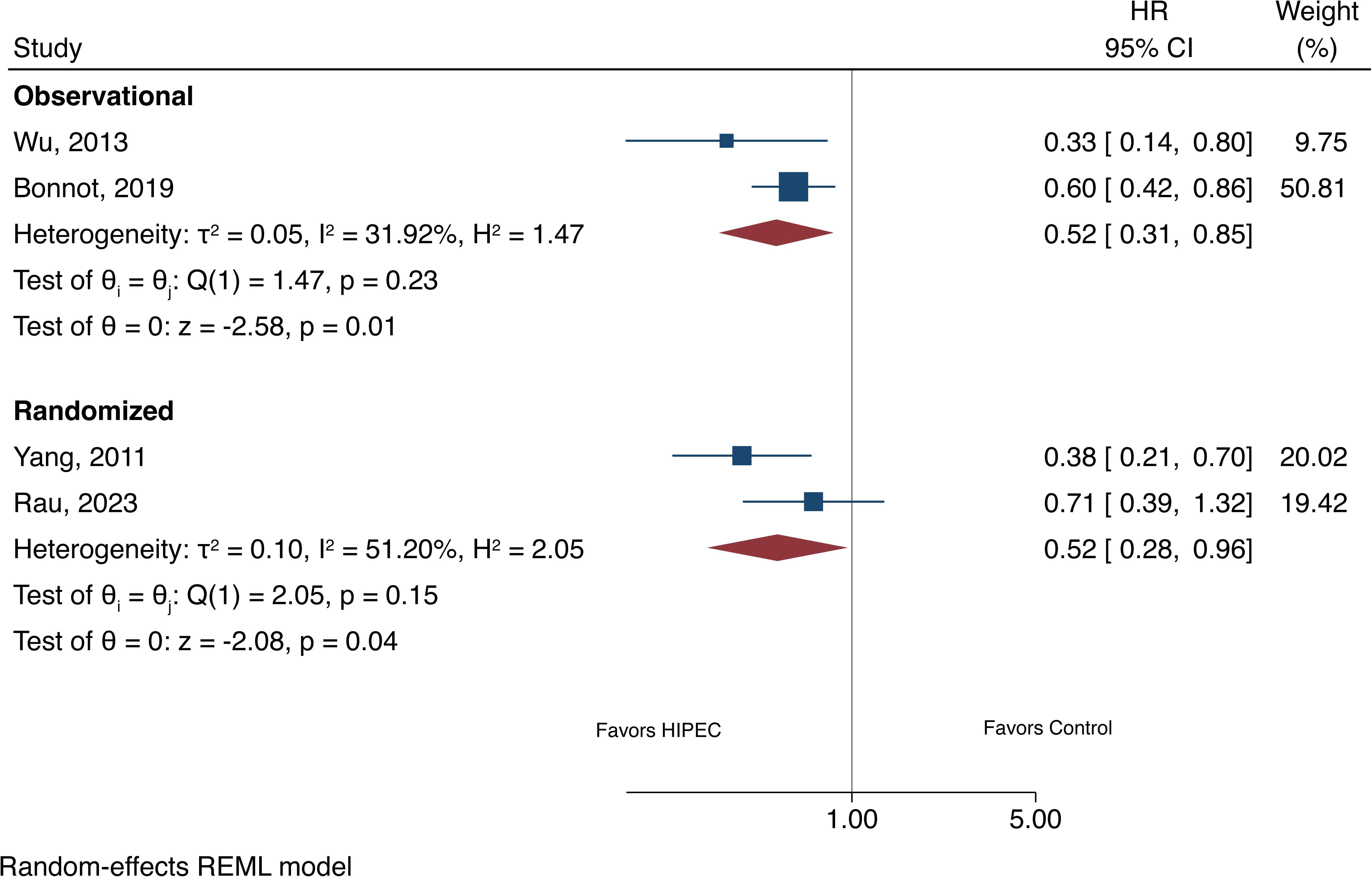
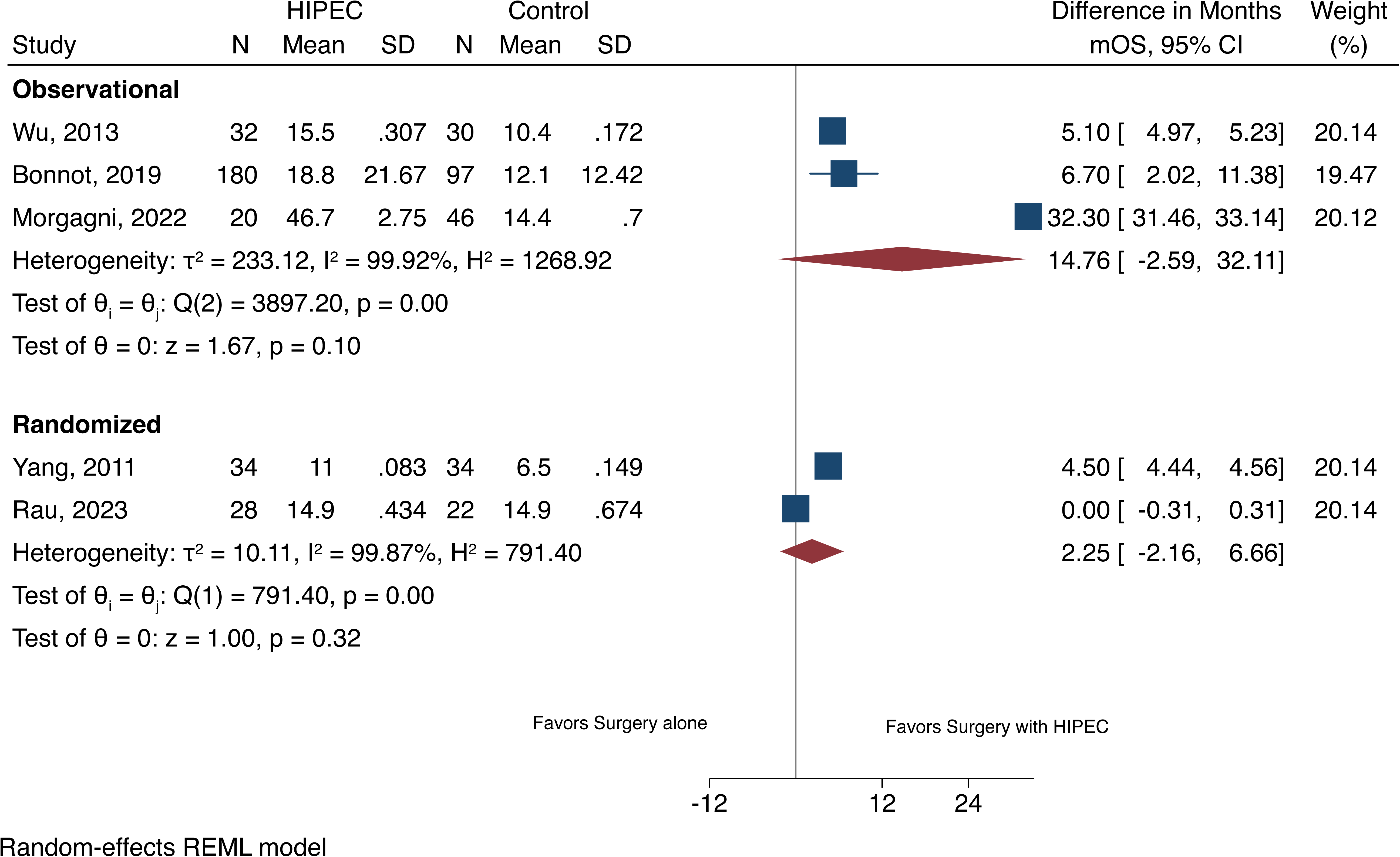
Meta-analysis for median overall survival comparing CRS-HIPEC to CRS alone

**Figure 4:**
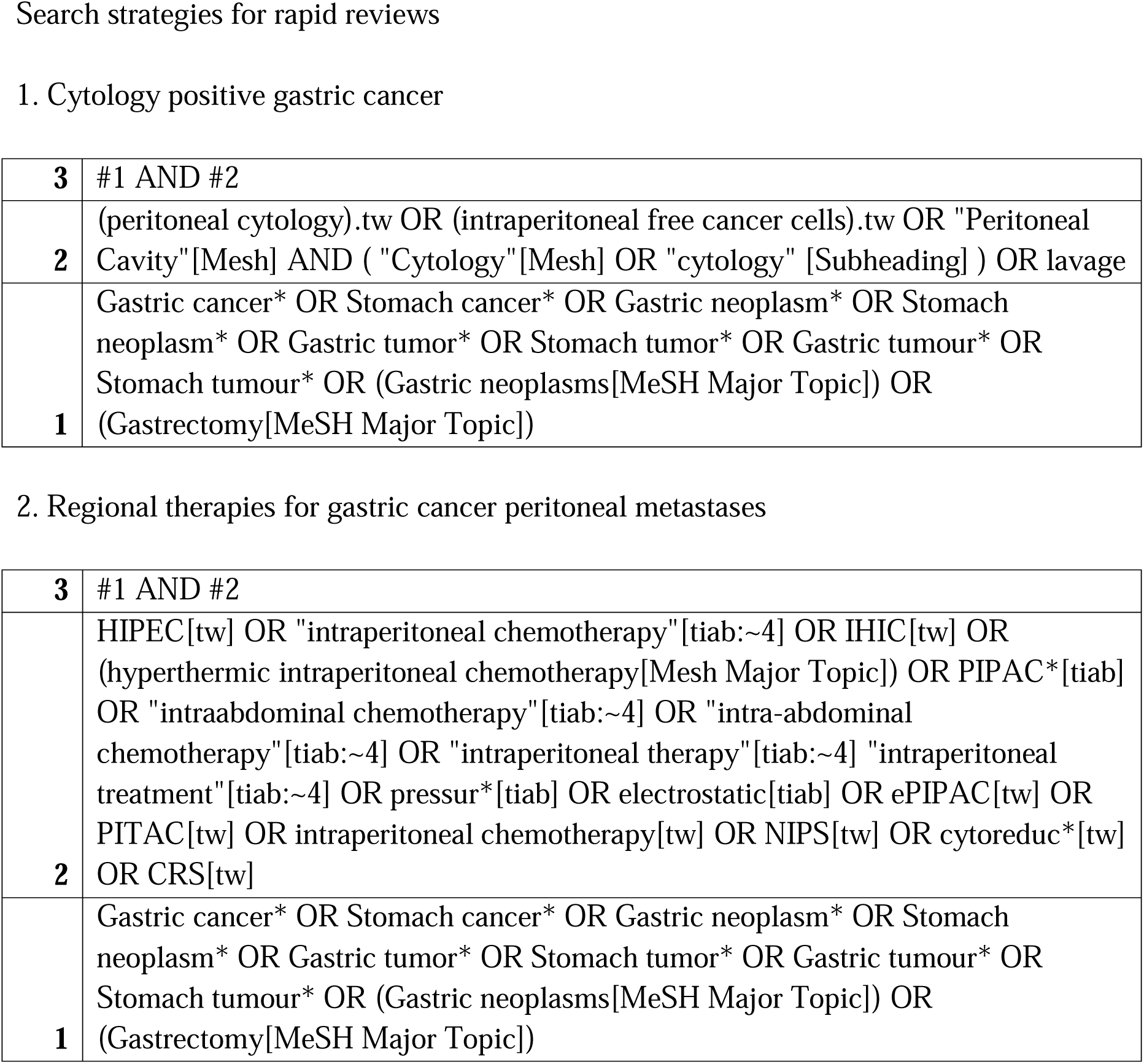
Search strategies for gastric cancer with peritoneal metastases

#### Block 6 (Agreement: Round 1 83%; Round 2 85%)

##### Recommendation

**In patients with PCI < 7-10 but in whom complete cytoreduction is not predicted, or their functional status would not permit an extensive surgery, intraperitoneal chemotherapy with or without additional systemic therapy may be considered.**

##### Port-Based Approaches

The implantation of a peritoneal port is considerably less invasive than HIPEC, allows for repeated IP administration of chemotherapy, and leads to high concentrations of chemotherapeutic drugs in the peritoneal cavity, allowing prolonged direct exposure of free cancer cells or peritoneal deposits. The only RCT that examined the role of IPC is the PHOENIX-GC trial, which found that there was no difference in survival between IPC plus systemic therapy versus systemic therapy alone.^54^ However, subsequent analyses that adjusted for baseline ascites showed potential benefits of the IP regimen. In addition, there was a crucial imbalance in the amount of ascites that favored the systemic therapy alone group.

Other observational studies have combined IP and IV chemotherapy with cytoreduction and HIPEC. This is referred to as Neoadjuvant Intraperitoneal-Systemic Chemotherapy (NIPS) or bidirectional therapy (BIPSC). Supplemental Table 8 shows the results of our rapid review for single-arm studies evaluating BIPSC. Notably, several groups have shown that BIPSC prolonged survival in patients with GCPM.^55,56^

Several groups have compared BIPSC to chemotherapy alone (see Supplemental Table 7). Kim et al. found that patients who underwent minimally invasive surgery following NIPS had a higher 2-year progression free survival rate than those who only underwent NIPS (36.4% vs. 10.5%); of note, this was a propensity weighted study.^57^ Lei et al. found that BIPSC had better overall survival (15.9 vs. 10.8 months) and 3-year overall survival rates (18.4% vs. 10.1%) as compared to chemotherapy alone.^58^ In 2016, Yuan et al. showed that BIPSC had better median overall survival (494 vs. 223 days) and progression free survival (164 vs. 129 days) compared to chemotherapy alone.^59^ Lee et al. found that patients who underwent L-HIPEC + NIPS followed by CRS-HIPEC had a better mean overall survival compared to those who only underwent CRS-HIPEC, chemotherapy or palliative care.^60^

##### Laparoscopic HIPEC (L-HIPEC)

Given the high morbidity associated with combining HIPEC and CRS, there has been an effort to administer HIPEC in a minimally invasive fashion, so as to decrease the associated morbidity. First published by Yonemura in 2016, other groups have shown that L-HIPEC was well tolerated and could reduce PCI score.^61,62^ Survival outcomes were examined by the Badgwell group of patients treated in with L-HIPEC and reported in 2020.^63^ They found that the median overall survival was 24.7 months in L-HIPEC and 21.3 months in standard care patients. Of note, almost all studies evaluating the efficacy of L-HIPEC exclude patients with high volume peritoneal disease. While a survival benefit of L-HIPEC has yet to be shown with small studies, larger more strongly powered RCTs are necessary.

##### Pressurized intraperitoneal aerosol chemotherapy (PIPAC)

PIPAC is a novel technique delivering drugs into the abdominal cavity as an aerosol under pressure. The theory behind PIPAC is that by creating an artificial pressure gradient within the intraperitoneal cavity, there will be enhanced tissue uptake and distribution of the aerosolized drug within the abdominal cavity. In 2016, Nadiradze preformed a retrospective analysis of 60 PIPAC procedures applied in 24 consecutive patients with GCPM (average PCI 16).^64^ They found that the median survival time was 15.4 months, and 9 patients had severe adverse events. Several other groups have discovered similar safety profiles of PIPAC (see Supplemental Table 1). PIPAC has also been incorporated into BIPSC. Most recently, Casella et al., showed that PIPAC used in a bidirectional approach is safe and feasible.^65^ A phase III trial labeled PIPAC VEROne by the same group will evaluate secondary resectability rate and survival statistics.

#### Block 7 (% Agreement: Round 1 91%; Round 2 96%)

CRS with IPCT is not recommended in patients with PCI > 7-10 despite stable/responsive disease. The survival benefit is reduced in these patients and a risk of substantial morbidity exists with the surgery and chemotherapy. Instead, these patients should be referred for further lines of systemic therapy, a clinical trial, or supportive care.

#### Block 8 (Agreement: Round 1 92%; Round 2 99%)

##### Recommendation

**Armed with the last 15 years of research, we worked with medical oncologists in the Gastric Cancer Working Group to create first-line systemic therapy recommendations (Table 3). For more detailed therapies, NCCN guidelines should be referenced.** If after diagnosis, patients are determined to have a high burden of disease on cross sectional imaging and/or laparoscopy, they should be referred for further lines of systemic therapy, a clinical trial, or supportive care. If they respond to further lines of systemic therapy, candidacy for regional therapeutic interventions may be re-assessed based on discussions with a multi-disciplinary team.

##### Systemic Therapy

There are several challenges with systemic therapies for the treatment of GCPM. The presence of the plasma-peritoneal barrier and the poor blood supply of peritoneal metastases limit the therapeutic effect of systemic agents. Additionally, patients with GCPM often develop complications such as poor nutrition and decreased performance status that hinder their ability to receive systemic therapy. The goals of palliative intent systemic therapy include delaying disease progression and increasing overall survival, controlling cancer-related symptoms and maintaining or improving quality of life. Several factors need to be considered when deciding on choices of systemic therapy including treatment goals, burden of disease, molecular characteristics, patients’ performance status, organ function and general tolerability to systemic therapy along with availability of treatment options. In general, GCPM patients are included in most systemic therapy trials for metastatic or stage IV gastric cancer. However, outcomes of GCPM compared to other sites of distant metastases such as the liver or para-aortic lymph nodes are not reported consistently as subgroup analyses. This has led to difficulty in discerning the benefit of these therapies in the context of GCPM specifically. Most of the trials that report GCPM subgroups show a similar or lower benefit for the intervention arm when compared to non-GCPM subgroups, reinforcing the principle of resistance of peritoneal metastases to systemic therapy. Nevertheless, several therapies have been well studied and approved for the treatment of GCPM.

###### First-line treatment: Platinum and Fluoropyrimidine based chemotherapy doublet

Chemotherapy has been shown to prolong survival and improve symptom control.^66^ The combination of fluoropyrimidine and platinum has been established as standard of care chemotherapy backbone for patients fit for doublet treatment.^67^ The REAL-2 study demonstrated interchangeability and non-inferiority between cisplatin and oxaliplatin, as well as capecitabine and infusional 5FU. In general, oxaliplatin is preferred due to better tolerance and side effect profile.^68^

###### Immune Checkpoint Inhibitors

CHECKMATE-649 and KEYNOTE-859 are randomized phase III trials that have demonstrated the benefit of the addition of nivolumab and pembrolizumab to chemotherapy in the first-line treatment of metastatic gastric cancer.^69,70^ Neither trial has reported GCPM specific outcomes. There remains much controversy on the role of PD-L1 as a biomarker for selecting for patients that may derive the maximum benefit from treatment with anti-PD-1 immune checkpoint inhibition and is beyond the scope of this article.^71^ The ATTRACTION-2 study demonstrated an overall survival benefit of nivolumab compared to placebo for patients with metastatic gastric cancer that had progressed on at least two prior lines of therapy (i.e. third line and beyond). The GCPM subgroup had a lower benefit from nivolumab treatment (HR 0.74, 95% CI: 0.48 – 1.15) compared to the non-GCPM subgroup (HR 0.64, 95% CI: 0.50 – 0.82).

###### Targeted therapy

Several targeted therapies have now been approved for the treatment of metastatic gastric cancer. Ramucirumab, an anti-angiogenic agent, has been approved in the second line, either as a single agent or in combination with paclitaxel chemotherapy, based on the REGARD and RAINBOW trials. In both trials, the GCPM subgroups appeared to benefit from ramucirumab treatment, but to a lesser extent compared to the non-GCPM subgroup.

The addition of trastuzumab to platinum-fluoropyrimidine doublet chemotherapy for HER2 positive gastric cancer was established as a standard-of-care based on the TOGA trial.^72^ More recently, the addition of pembrolizumab to the combination of trastuzumab and chemotherapy was shown to have an improvement in survival in the KENOTE-811 trial, particularly in the PD-L1 positive subgroup, and has attained regulatory approval.^73^ However, neither of the trials report GCPM subgroup outcomes.

More recently, the addition of Zolbetuximab to chemotherapy in CLDN18.2 positive metastatic gastric cancer has demonstrated a survival benefit and is pending regulatory approval. GCPM subgroup data has not been reported to date.

## DISCUSSION

Herein we summarize the updated consensus guidelines on the management of gastric cancer with PMs. Our current consensus group expanded to include surgical oncologists, medical oncologists, radiologists, pathologists, and patient advocates. Consensus was achieved in all eight question blocks after two rounds of review. Despite the low to moderate level of evidence, substantial work had been produced in the field of GCPMs to require major adoptions and revisions.

There were two areas of contention within the care pathway, namely blocks five and six. The REGATTA trial provided negative results on the value of CRS-HIPEC compared to chemotherapy alone, while the GYMSSA trial supported the use of CRS-HIPEC over chemotherapy. However, given the numerous observational studies that have shown the benefit of CRS-HIPEC over chemotherapy, the disease site working group felt strongly that it should be recommended in patients with whom a CC-0 resection is predicted.

Additionally, the two RCTs comparing CRS-IPC to CRSa also contradict each other. Yang et al. showed that median overall survival was improved with the addition of IPC to CRS, while GASRIPEC-1 did not show any difference between the two groups. Again, four observational studies have shown that the addition of CRS to IPC does improve survival. With this information, the disease site working group felt strongly that the addition of IPC has value for GCPM patients when combined with CRS.

As for block 6, the PHOENIX-GC trial remains the only RCT examining the utility of BIPSC alone compared to systemic therapy, and it failed to show a survival benefit for GCPM patients. Several observational studies afterwards have shown that BIPSC in addition to CRS-HIPEC does provide a survival benefit. Furthermore, L-HIPEC and PIPAC remain safe and feasible options for GCPM patients, especially in converting patients with a high burden of disease to a level acceptable for a CC-0 resection. Therefore, we recommend using these treatment modalities in conjunction with CRS and IPC in patients with a low PCI and a level of disease that may preclude them from moving directly to CRS.

Major limitations of this expert consensus merit discussion. Firstly, the expert panel consisted primarily of surgical oncologists. Having expected this bias from the inception phases, thought leaders in medical oncology and other disciplines were involved early on for reviewing feedback from the first Delphi round and outlining principles of systemic therapy. Secondly, the Delphi consensus entailed voting on blocks rather than individual itemized recommendations, aligning with the original Chicago Consensus framework. While this approach helped mitigate survey fatigue, it may have may have compromised the granularity of feedback received. Finally, there were one or two members engaged at each level of the rapid review process, but many more were only involved for one or two stages. This could have led to different interpretations of the criteria used to screen literature and extract data. The two-person verification system should have mitigated this effect, however.

### International Perspective

There are several notable international guidelines for management of gastric cancer, ranging from individual countries to large multi-national organizations.^74–78^ All of these guidelines recommend palliation in the form of supportive care and systemic chemotherapy for GCPM. However, it is worthwhile examining these guidelines’ recommendations for staging laparoscopy and surgery in the management of GCPM.

With regard to staging laparoscopy, the 2018 Korean national guidelines recommend peritoneal washing cytology for all patients, given that cytology positive patients are associated with cancer recurrence and poor prognosis. The 2016 PSOGI guidelines also recommend staging laparoscopy in all patients with gastric cancer. In 2020, the French Association of Surgery disagreed with this consensus, recommending that exploratory laparoscopy only be carried out only in patients with cT3/T4 and/or N + disease. The European Society for Medical Oncology (ESMO) and pan-Asian adapted ESMO guidelines agreed with this narrowing of criteria, recommending diagnostic laparoscopy and peritoneal washings for cytology only in selected patients with resectable gastric cancer.

Additionally, surgery for GCPM has been controversial. Both ESMO and the pan-Asian guidelines reference the Phase III REGATTA trial to recommend against gastrectomy in these patients. The caveat to this is that they recommend the resection of metastases on an individual basis, especially those who respond to chemotherapy. This contrasts the 2016 PSOGI guidelines, which suggests that CRS combined with perioperative intraperitoneal/systemic chemotherapy is the only strategy to improve the long-term survival of GCPM patients. They do note, however, that CRS should be offered in patients with low PCI level and negative cytology. The French guidelines echo this sentiment, adding a PCI cut-off of 7 and implementing HIPEC alongside CRS. Chinese guidelines only recommend “reductive surgery” for patients with GCPM and urgent symptoms, such as bleeding or obstruction. Finally, Korean guidelines suggest that CRS can be considered for locally advanced unresectable or cM1 gastric cancer not detected in preoperative evaluation but incidentally identified during surgery and if R0 resection is possible.

In patients with a high PCI, conversion therapy and subsequent surgery is also highly debated. PSOGI recommends reducing PCI with neoadjuvant L-HIPEC and/or bidirectional therapy. They do not comment on subsequent surgery, though. In China and Korea, if systemic chemotherapy leads to complete resolution of PM, conversion gastrectomy is recommended. For French centers, GCPM patients with high PCI should undergo IV chemotherapy or PIPAC alternating with IV chemotherapy. They do note that this is an expert opinion not based on strong evidence. ESMO and pan-Asian adapted ESMO do not make recommendations on patients who are successfully converted to negative cytology and/or lower PCIs.

### Patient/Caregiver Perspective

Understanding the impact of GCPM on patients and their caregivers is crucial to their holistic treatment. Organizations like Hope for Stomach Cancer (STOCAN) offer invaluable resources, fostering early detection, clinical trial access, and a supportive community for patients. Through STOCAN, we were able to connect with patients about their experiences with clinical trials and research in the GCPM space. For many patients, enrollment in trials instills hope, offering not only potential survival benefits but also a sense of purpose through contributing to research. Although clinical trial availability may be ample, navigating enrollment often requires tenacity and robust support networks. Patients prioritized overall survival as a primary outcome measure for research, while also valuing progression-free survival and recurrence rates. They emphasized the importance of incorporating quality-of-life metrics into outcome measures. Additionally, patients highlighted the necessity of diverse support networks, blending online and offline resources, including friends, family, peers, and medical professionals. The medical team’s guidance is vital, directing patients to specialized centers when necessary. Overall, patient perspectives underscore the significance of holistic care and collaborative support networks in navigating the challenges of GCPM management.

## CONCLUSION

In summary, we reported an updated Delphi consensus on the management of GCPM that included a multidisciplinary team of experts. Preoperative evaluation should be comprehensive and include genetic testing and a diagnostic laparoscopy to assess peritoneal disease burden. In patients with high PCI (> 10), supportive care should be offered, whereas in patients with low PCI, they should be enrolled in systemic therapy based on their genetic status. In those that respond to systemic therapy, we recommend CRS-HIPEC for patients with predicted CC0 or BIPSC prior to CRS-HIPEC. Finally, in those that continue to have a high PCI after therapy, systemic therapy and/or a clinical trial is recommended.

## Data Availability

The datasets used and/or analyzed during the current study are available from the corresponding author on reasonable request.

---

**Supplemental Table 1:** Summary of landmark PIPAC trials in the past 10 years.

| Study | Intervention | Intraperitoneal chemotherapy | Systemic chemotherapy | Overall survival (median, months) | Adverse events |
| --- | --- | --- | --- | --- | --- |
| Casella 2023<sup>1</sup> | Bidirectional | First 29 procedures: Cisplatin at 7.5 mg/m<sup>2</sup> + Doxorubicin at 1.5 mg/m<sup>2</sup><br>Subsequently: Cisplatin at 10.5 mg/m<sup>2</sup> and doxorubicin at 2.1 mg/m<sup>2</sup>. Done at a constant capnoperitoneum of 12 mmHg at 37 °C for 30 minutes. | For most patients, ST given before or after PIPAC. (Regimens variable) | 10.5 | CTCAE 3/4: 2 patients<br>CTCAE 5: None |
| Sindayigaya 2022<sup>2</sup> | PIPAC | Cisplatin at 7.5 mg/m<sup>2</sup> + Doxorubicin at 1.5 mg/m<sup>2</sup>. Done at a constant capnoperitoneum of 12 mmHg at 37 °C for 30 minutes. | Concurrent systemic chemotherapy given in 34 (23.8%) patients / 194 (65.5%) PIPAC procedures. (Regimens not specified) | 11 | CTCAE 3: 7 patients<br>CTCAE 4: None<br>CTCAE 5: 2 patients died |
| Alyami 2021<sup>3</sup> | Bidirectional | Cisplatin at 7.5 mg/m<sup>2</sup> + Doxorubicin at 1.5 mg/m<sup>2</sup>. Done for 30 minutes. | Yes (Regimens not specified) | 19.1 | CTCAE 3: 10 procedures<br>CTCAE 4: 5 procedures<br>CTCAE 5: 2 patients died |
| Ellebæk 2020<sup>4</sup> | PIPAC | Cisplatin at 7.5 mg/m<sup>2</sup> + Doxorubicin at 1.5 mg/m<sup>2</sup>. Done for 30 minutes. (7 patients had an additional ePIPAC step after PIPAC delivery) | 9/20 received systemic chemotherapy. (Regimens not specified) | 4.7 | CTCAE 3: 1 patient<br>CTCAE 4: 1 patient<br>CTCAE 5: - |
| Struller 2019<sup>5</sup> | PIPAC | Cisplatin at 7.5 mg/m<sup>2</sup> + Doxorubicin at 1.5 mg/m<sup>2</sup>. Done at a constant capnoperitoneum of 12 mmHg at 37 °C for 30 minutes. | PIPAC alone (No systemic chemotherapy allowed) | 6.7 | CTCAE 3: 3 patients<br>CTCAE 4: None<br>CTCAE 5: None |
| Gockel 2018<sup>6</sup> | PIPAC | Cisplatin at 7.5 mg/m<sup>2</sup> + Doxorubicin at 1.5 mg/m<sup>2</sup>. Done at a constant capnoperitoneum of 12 mmHg at 37 °C for 30 minutes. | Yes (Regimen not specified) | 4 | CTCAE 3: None<br>CTCAE 4: None<br>CTCAE 5: None |
| Khomyakov 2016<sup>7</sup> | Bidirectional | Cisplatin at 7.5 mg/m<sup>2</sup> + Doxorubicin at 1.5 mg/m<sup>2</sup>. Done at a constant capnoperitoneum of 12 mmHg at 37 °C for 30 minutes. | Yes (XELOX)- 3 to 4 courses | 13 | CTCAE 3: -<br>CTCAE 4: None<br>CTCAE 5: None |
| Nadiradze 2016<sup>8</sup> | PIPAC | Cisplatin at 7.5 mg/m<sup>2</sup> + Doxorubicin at 1.5 mg/m<sup>2</sup>. Done at a constant capnoperitoneum of 12 mmHg at 37 °C for 30 minutes. | 8/24 (33%) received systemic chemotherapy. (Regimens not specified) | 15.4 | CTCAE 3: 6 patients<br>CTCAE 4: 1 patient<br>CTCAE 5: 2 patients |
| Tidadini 2022<sup>9</sup> | Bidirectional vs Systemic Therapy (ST) alone | Cisplatin at 7.5 mg/m<sup>2</sup> + Doxorubicin at 1.5 mg/m<sup>2</sup>. In the case of contraindication, oxaliplatin at a dose of 92 mg/m<sup>2</sup> was used. Done at a constant capnoperitoneum of 12 mmHg at 37 °C for 30 minutes. | Yes (Regimens variable) | PIPAC+ST: 12.8<br>ST alone: 9.1 | CTCAE not reported;<br>Clavien-dindo 3b/4 in:<br>PIPAC+ST: 2/17 (11.8%)<br>ST alone: 2/29 (6.9%) |

**Supplemental Table 2:** Summary of studies included in KQ 1, particularly patients who converted from cytology positive to negative.

| Study | Type | Duration | Sample Size | Intervention | Survival | Morbidity/Mortality | Notes |
| --- | --- | --- | --- | --- | --- | --- | --- |
| Emoto 2015<sup>10</sup><br>*Japan* | Retrospective Cohort | 2005 – 2012 | CY1 45<br>CY0 9 | Oral S-1 and Paclitaxel IV and IP. IP given concurrently with IV infusion. | Converted negative MST 20.0 months<br>Not converted MST 13.0 months | Not stated | None |
| Lorenzen 2010<sup>11</sup><br>*Germany* | Retrospective Cohort | 1994 – 2000 | CY1 22<br>CY0 39 | Cisplatin, 5-FU (PLF). Surgery performed if cytology converted to negative. | Converted negative MST 36.1 months, 2- and 5-year was 71.4<br>Not converted MST 9.2 months; 2-year 25%, 5-year 8.3% | Not stated | None |
| Yago 2022<sup>12</sup><br>*Japan* | Retrospective Cohort | 2006 – 2019 | Surgery First: 21<br>Chemo First: 17 | Adjuvant Oral S-1. Surgery performed if cytology converted to negative. | Converted negative MST not reached; 3-year 75%<br>Not converted 10.9 months, 3-year 0% | 3-year RFS: Surgery first 17.1%; Chemo first 17.4% | 32 recurrences; 78% peritoneal |
| Yamaguchi 2021<sup>13</sup><br>*Japan* | Retrospective Cohort | 2008 – 2012 | Surgery First: 563<br>Chemo First: 150 | Chemotherapy not specified. Surgery performed if cytology converted to negative. | Converted negative MST 32 months<br>Not converted MST 18.8 months | Median PFS: Chemo 14.9 months; Surgery: 13.9 months | Median OS: Chemo 24.8 months; Surgery 24.0 months |
| Yamamoto 2015<sup>14</sup><br>*Japan* | Retrospective Cohort | 1999 – 2014 | Neoadjuvant 10<br>Adjuvant 13 | Neoadjuvant regimens varied. Adjuvant with oral S-1. Surgery performed if converted negative with neoadjuvant chemotherapy. | Converted negative MST 26.5 months<br>Not converted MST 13.6 months | Neoadjuvant: MST 24.7 months; 5-year 15% \| Adjuvant: MST 17.1 months; 5-year 15% | 8/23 underwent surgery after conversion |
| Yasufuku 2020<sup>15</sup><br>*Japan* | Retrospective Cohort | 2009 – 2015 | Conversion: 13<br>Palliative: 19 | Chemotherapy varied based on HER2 status. Postop chemo was S-1 for 1-year after R0 resection and until progression in R1. | Converted Negative MST 108.5 months, 3-year survival was 76.9%<br>Not converted MST 16.3 months, 3-year survival was 10.5% | Overall MST 24.1 months<br>PFS: Conversion 27.9 months; Palliative 8.1 months | None |
| Valletti 2021<sup>16</sup><br>*Switzerland* | Retrospective Cohort | 2011 – 2019 | CY0: 125<br>Converted: 9<br>CY1: 5<br>PC: 33 | NACT via FLOT or EF. Surgery performed 3-4 weeks after NACT if converted. | Converted Negative MST not reached, 3-year survival 65%<br>Not Converted: 13 months, 3-year survival 0% | DFS: Converted: 21.7 months; 1-year 64% \| Not converted: 1-year 20% | None |

**Supplemental Table 3:** Summary of studies included in KQ1, particularly those that directly compared cytology positive to negative.

| Study | Type | Duration | Sample Size | Intervention | Survival | Morbidity/Mortality | Notes |
| --- | --- | --- | --- | --- | --- | --- | --- |
| Masuda 2015<sup>17</sup><br>*Japan* | Retrospective Cohort | 1995 – 2005 | P0CY0: 23<br>P0CY1: 37 | All patients received extensive intraperitoneal lavage (EIPL). Then IPC (Cisplatin) followed by adjuvant therapy (5-FU) | CY0: 5-year survival 33.9%<br>CY1: 5-year survival 46.5% | Recurrences: 19 (51.4%) in CY1 and 13 (56.5%) in CY0 | CY0 is Stage II B+C GC |
| Yamaguchi 2020<sup>18</sup><br>*Japan* | Retrospective Cohort | 2007 – 2012 | P0CY1: 367<br>P1aCY0: 81<br>P1aCY1: 58 | Surgical resection followed by post operative chemo was generally 6 weeks of S-; CS comprised of oral S-1 then IV cisplatin. | <u>Mean Overall Survival</u><br>**S1 Chemo:** P0CY1, P1aCY0, and P1aCY1 were 30.7, 26.9, and 22.9 months<br>**S1 + Cisplatin:** 24.4, 25.3, and 24.7 months<br>**Other chemo:** 24.1, 26.5, and 39.2 months' | <u>5-Year Survival</u><br>**S1 Chemo:** P0CY1, P1aCY0, and P1aCY were 30.2, 19.9, 17.5%<br>**S1 + Cisplatin:** P0CY1, P1aCY0, and P1aCY were 26.8, 17.1, and 11.1%<br>**Other chemo:** P0CY1, P1aCY0, and P1aCY were 20.9, 33.3 and 16.9% | None |
| Fukagawa 2010<sup>19</sup><br>*Japan* | Retrospective Cohort | 1992 – 1993 | P0CY0: 485<br>P0CY1: 88<br>P1CY0: 22<br>P1CY1: 106 | Surgical resection followed by post-operative adjuvant chemotherapy, fluorouracil-based | P0CY1 MST 12 months; 2-year and 5-year were 25.3% and 7.8% | P0Cy1, type 4 gastric cancer had worst survival. MST 10 months. | Contains patients who had locally resectable PC and those with diffuse dissemination detected at surgery |
| Kanazawa 2013<sup>20</sup><br>*Japan* | Prospective, non-randomized, open-label clinical trial | 2006 – 2010 | P0CY1: 8<br>P1CY0: 14<br>P1CY1: 6 | Gastrectomy without preoperative chemotherapy. Then oral S-1 and IV docetaxel. | P0CY1: MST 34.5 months<br>P1CY0: MST 34.3 months<br>P1CY1: MST 19.3 months | Median PFS was 22.9 months | None |

**Supplemental Table 4:** Summary of outcomes in KQ1 for comparative and single-arm studies evaluating surgery in cytology positive patients.

| Study | Type | Duration | Sample Size | Intervention | Survival | Morbidity/Mortality | Notes |
| --- | --- | --- | --- | --- | --- | --- | --- |
| Kang 2021<sup>21</sup><br>*Japan* | Retrospective Cohort | 2013 – 2018 | POCY1: 48 | Radical gastrectomy performed on all patients. 12 received NACT, 30 received adjuvant, 3 received adjuvant chemoradiotherapy | MST 22 months; 1-, 2-, 3- and 5-year survival rates were 72.4%, 47.8%, 32.9%, and 20.5% | Not stated | None |
| Kuramoto 2009<sup>**22</sup><br>*Japan* | Randomized controlled trial | 1995 – 2005 | EIPL-IPC: 30<br>IPC: 29<br>Surgery Alone: 29 | All patients received adjuvant chemo via 5 FU derivatives for 2 years. | EIPL-IPC: MST 35 months, 5-year 43.8%<br>IPC: MST 16 months, 5-year 4.6%<br>Surgery Alone: MST 15 months, 5-year 0% | Peritoneal recurrence rate was 69.3% | None |
| Oh 2012<sup>23</sup><br>*Korea* | Retrospective Cohort | 1994 – 2006 | POCY1: 37 | Curative resection followed by adjuvant therapy (22/37), chemoradiotherapy (9/37), or no adjuvant therapy. Chemotherapy varied. | Mean and median OS were 15.5 (9.5) and 18 months (2-45) respectively. 1-, 3-, and 5-year survivals were 43.2%, 5.4%, and 0% | Mean and Median DFS were 10.7 (7.5) and 10 months (1-33), respectively | All patients recurred within 3 years. |
| Shimada 2002<sup>24</sup><br>*Japan* | Retrospective Cohort | 1989 – 2009 | Surgery (S): 8<br>S + IPC: 7<br>S + EIPL-IPC: 7 | Curative resection followed by no additional therapy; IPC was performed with Cisplatin (100 mg). Chemo varied. | S: 2-year 0%<br>S + IPC: 2-year 15.3%<br>S + EIPL-IPC: 2-year survival 57.1% | Peritoneal Recurrence: S (100%), S + IPC (85.7%), S + EIPL-IPC (42.9%) | No demographic information in this study. |
| Shim 2020<sup>25</sup><br>*Korea* | Retrospective Cohort | 2004 – 2016 | Chemo + Surgery: 64<br>Surgery Alone: 24 | Curative resection or resection plus adjuvant chemotherapy with TS-1, XELOX, FOLFOX, Cisplatin + TS-1 (CS) | Chemo: MST 25.5 months; 1-year 88.7%<br>Surgery alone: MST 12.1 months; 1-year 50% | Chemo: Median DFS 11.63; 1-year DFS 46.9% \| Surgery: Median DFS 6.98; 1-year DFS 12.5% | None. |
| Yamanaka 2013<sup>26</sup><br>*Japan* | Retrospective Cohort | 2000 – 2010 | Open: 9<br>Laparoscopic: 9 | Patients received open or laparoscopic cytoreductive surgery. If patients were ECOG 0-1, they received adjuvant chemotherapy via S-1 80 mg/m<sup>2</sup> or IV 500 mg/m<sup>2</sup> 5-FU + cisplatin | Open: MST 13.1 months; 1- and 2-year survival rates were 50 and 33.3%<br>Lap: MST not reached; 1- and 2- year survival rates were 100 and 62.5% | Quality of surgery did not differ. Lap had lower EBL, less LOS, and quicker resumption of food intake. | None. |
| Kobayashi 2021<sup>27</sup><br>*Japan* | Retrospective Cohort | 2008 – 2015 | Surgery: 74<br>No primary surgery: 30 | 5 patients received neoadjuvant chemotherapy, while 82 received some form of adjuvant chemotherapy | Gastrectomy: MST 21.7 months<br>Non-surgery: MST 20.5 months | Chemo MST 23 vs. non-chemo 8.6 months | None. |
| Kano 2017<sup>28</sup><br>*Japan* | Retrospective Cohort | 2000 – 2015 | No chemo: 9<br>Chemo: 36 | Macroscopic resection followed by post operative S1 4 weeks followed by 2 weeks (1 course) or for 2 weeks followed by 1 week of rest (1 course). | Surgery Alone: MST 11.8 months, 1-, 3-, and 5-year OS were 44.4, 0.0, and 0.0%<br>Surgery + Chemo: MST 22.3 months, 1-, 3-, and 5-year OS were 83.3, 25.0 and 17.8% | Median RFS: Pathologic N0-3a 16.4 months; N3b 7.8 months | None. |
| Endo 2021<sup>29</sup><br>*Japan* | Retrospective Cohort | 2000 – 2019 | Total: 128 | Surgery in all patients. Preop chemo for 16 patients; post op chemo for 92 patients. Chemo varied widely. | MST 18.6 months \| 5-year survival: 19.6% | PFS: Median 12.9 months; 5-year 13.4% | None |
| Imano 2011<sup>30</sup><br>*Japan* | Retrospective Cohort | 2004 – 2009 | Surgery + IPC: 10<br>Controls: Unknown | IPC via paclitaxel. 9 patients received systemic chemo (S-1). Controls were historic CY1 patients who did not undergo any treatment | Surgery + IPC: MST 1151d; 2-year 70%; 3-year 56%<br>Control: MST 392 days; 2-year and 3-year 0% | Not stated. | No information on controls. |

**Supplemental Table 5:**
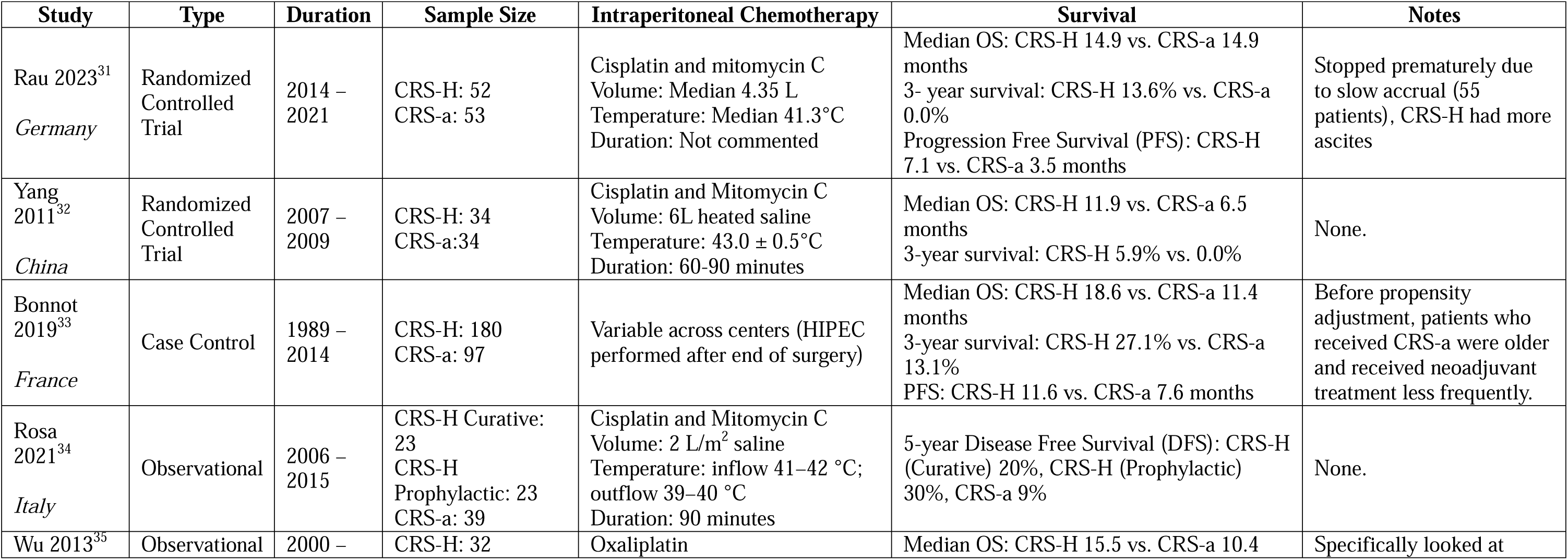
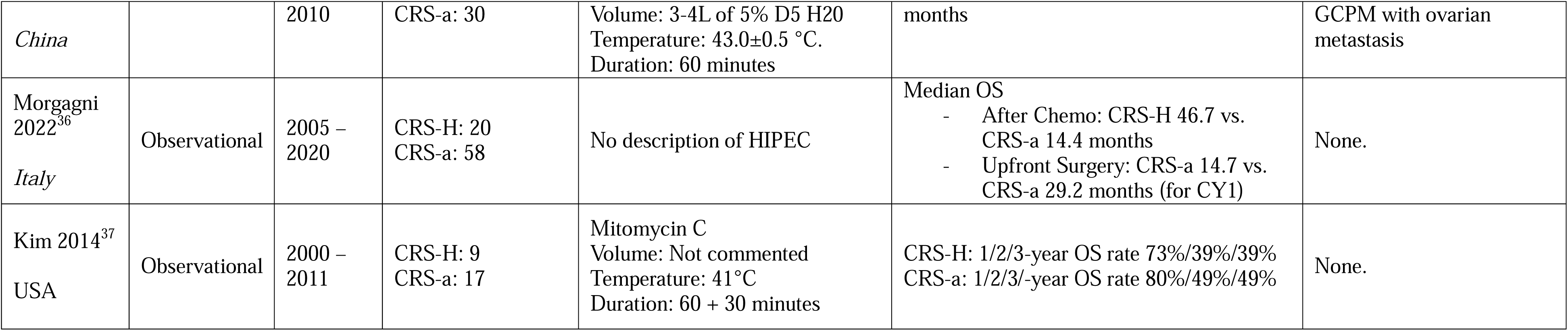
Summary of outcomes in KQ2 for comparative studies evaluating CRS-HIPEC vs. CRS-alone.

**Supplemental Table 6:**
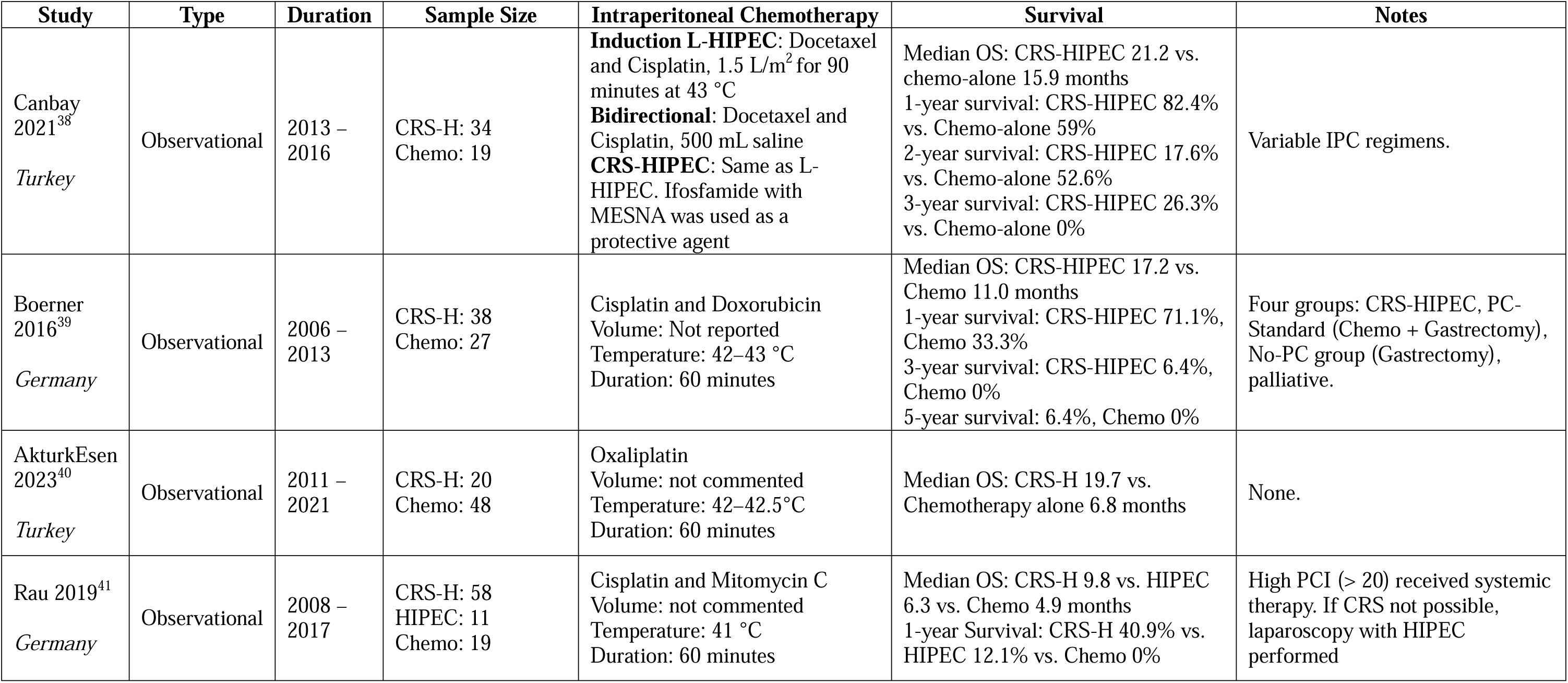
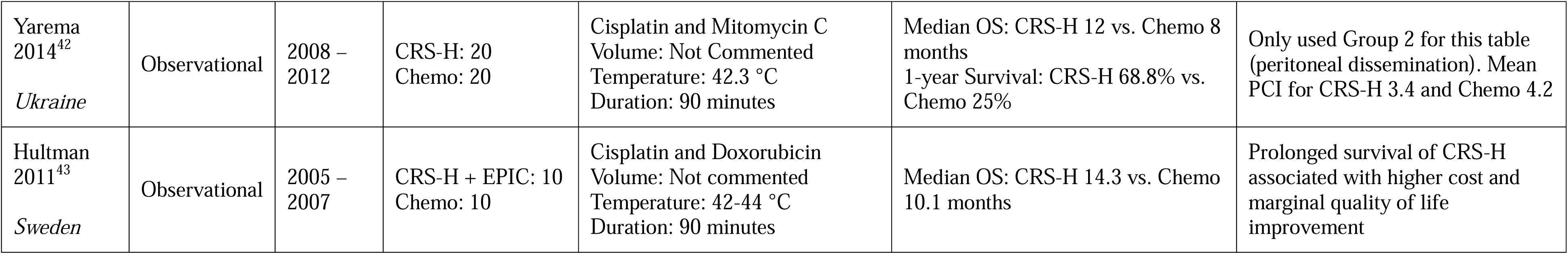
Summary of outcomes in KQ2 for comparative studies evaluating CRS-HIPEC vs. chemotherapy alone.

**Supplemental Table 7:** Summary of outcomes in KQ2 for comparative studies evaluating bidirectional therapy vs. chemotherapy alone.

| Study | Type | Duration | Sample Size | Intervention | Survival | Notes |
| --- | --- | --- | --- | --- | --- | --- |
| Kim 2022<sup>44</sup><br>*Korea* | Observational | 2011 – 2021 | Bidirectional: 26<br>Chemo: 26 | IP Paclitaxel was given alongside XELOX or FOLFOX, in 0.5 L NS over 2 hours. | Median PFS: Minimally Invasive Surgery + IPC (MIS-IPC) 13 months vs. systemic chemotherapy (SC) 6 months<br>2-year PFS: MIS-IPC 36.4% vs. SC 10.5% | Propensity weighted study. |
| Lei 2020<sup>45</sup><br>*China* | Observational | 2010 – 2017 | Bidirectional: 405<br>Chemo: 258 | IP Paclitaxel, Oxaliplatin or Cisplatin given in 2 L/m<sup>2</sup> at 43 +/- 0.1°C for 60 minutes. Systemic chemo was 5-FU or Paclitaxel based. | Median OS: Bidirectional 15.9 months vs. Chemotherapy only 10.8 months<br>3-year OS: Bidirectional 18.4% vs. Chemotherapy alone 10.1% | Bidirectional were younger and had more ascites. Thus, propensity score matching was used. |
| Yuan 2016<sup>46</sup><br>*China* | Observational | 2008 – 2014 | Bidirectional: 23<br>Chemo: 31 | Cisplatin given during chemotherapy. Details regarding HIPEC are missing. | Median OS: Bidirectional 494d vs. Chemo alone 223d<br>1-year survival: Bidirectional 41.7% vs. Chemo alone 23.8%<br>PFS: Bidirectional 164d vs. Chemo alone 129d | None. |
| Lee 2021<sup>47</sup><br>*Taiwan* | Observational | 2012 – 2020 | Conversion: 34<br>CRS-HIPEC: 15<br>Chemo: 23<br>Palliative: 23 | <u>If PCI > 12, L-HIPEC performed:</u> IP Oxaliplatin, Mitomycin C and Paclitaxel used in 2.5 L NS at 42°C for 90 minutes.<br><u>Bidirectional:</u> IP Paclitaxel + IV XELOX | Median OS: Conversion 18.8 months, CRS-HIPEC 13 months, chemo alone 8.3 months, palliative 5 months.<br>CC-0 MST 18.8 months vs. CC-1 10 months vs. CC-2 8.3 months | None. |
| Blumenthaler 2020<sup>48</sup><br>*USA* | Observational | 2013 – 2018 | L-HIPEC: 25<br>Chemo: 27 | Laparoscopic Cisplatin and Mitomycin C in 3-7L at 41-42°C inflow, 39-40°C outflow for 60 minutes | Median OS: Bidirectional 24.7 vs. Chemo-alone 21.3 months<br>1-, 2-, and 3-year OS rates: Bidirectional 95.5%, 57.2%, and 19.1% vs. Chemo-alone 76.9%, 19.1%, and 9.6% | None. |

**Supplemental Table 8:** Summary of outcomes for studies analyzed in KQ2 evaluating single arm bidirectional studies.

| Study | Type | Duration | Sample Size | Intervention | Survival | Notes |
| --- | --- | --- | --- | --- | --- | --- |
| Takeshita 2017<sup>49</sup><br>*Japan* | Observational | 2014 – 2015 | 19 | Laparoscopic Docetaxel and Cisplatin in 4L NS at 43°C for 40 minutes. Chemo via IV Docetaxel and Cisplatin, oral S-1. | Overall, 1-year OS 52.1%<br>CC0: 1-year OS 65.8%<br>CC1: 1-year OS 0.0% | None. |
| Badgwell 2017<sup>50</sup><br>*USA* | Phase II Trial | 2014 – 2016 | 19 | Laparoscopic Cisplatin and Mitomycin C in 1.5 L/min at 41-42°C for 60 minutes. Chemo regimens varied. | Median OS: 20.3 months<br>1- and 2-year OS 73.3% and 31.4% | None. |
| Canbay 2014<sup>51</sup><br>*Japan* | Observational | 2005 – 2012 | 194 | IP Docetaxel and Cisplatin and oral S-1. | \NIPS + CRS-HIPEC: 1-, 2- and 5-year OS - 62.7%, 29.5%, and 9.3%. NIPS with no response, CY1 or PC: 1-, 2- and 5-year OS 34%, 13.5%, and 0%. | None. |
| Yonemura 2012<sup>52</sup><br>*Japan* | Observational | 2004 – 2011 | 96 | IP Taxotere and Cisplatin in 0.5L NS and oral S-1. | CRS: Median OS 14.4 months, 1-, 3-, and 5-year OS 61%, 16%, and 16%<br>No CRS: 9.9 months, 5-y OS 0% | None. |
| Fujiwara 2012<sup>53</sup><br>*Japan* | Observational | 2006 – 2010 | 18 | IP Docetaxel in 1L NS with oral S-1. | Median OS: 24.6 months<br>1- and 2-year OS: 76% and 54% | None. |
| Yonemura 2009<sup>54</sup><br>*Japan* | Observational | 2002 – 2008 | 79 | IP Docetaxel and Cisplatin in 0.5L NS and oral S-1. | CRS: Median OS 1.7 years, 1-, 2-, and 3-year OS 67.4%, 40.0%, and 30.0%<br>No CRS: Median OS 0.88 years, 1-, 2-, and 3-year OS 35.9%, 20.4%, and not reported. | Positive cytology changed to be negative in 41 (63.0%) patients after NIPS. |
| Yonemura 2006<sup>55</sup><br>*Japan, USA* | Phase II Trial | 2001 – 2003 | 61 | IP Taxotere and Carboplatin in 1L NS for 30 minutes. Chemo via IV Methotrexate and 5-FU. | Median OS 14.4 months<br>1-year OS 67%. | None. |
| Ni 2017<sup>56</sup><br>*China* | Observational | 2010 – 2014 | 41 | IP Cisplatin at 43 +/- 0.5 °C for 120 minutes. Chemo via IV Docetaxel. | Median OS 8.6 months, 1-year OS: 24.4%<br>Complete response, partial response, non-partial response: Mean OS 15.4±5.6, 9.1±0.9, and 6.9±0.5 months, respectively | None. |
| Fujiwara 2011<sup>57</sup><br>*Japan* | Observational | 2000 – 2006 | 25 | IP Cisplatin and Mitomycin C. IV Docetaxel, 5-FU, and Cisplatin | Median OS 16.7 months<br>Median OS CY0 + PC reduced vs. CY1 or PC: 27.1 vs. 9.6 months | None. |

**Supplemental Table 9:**
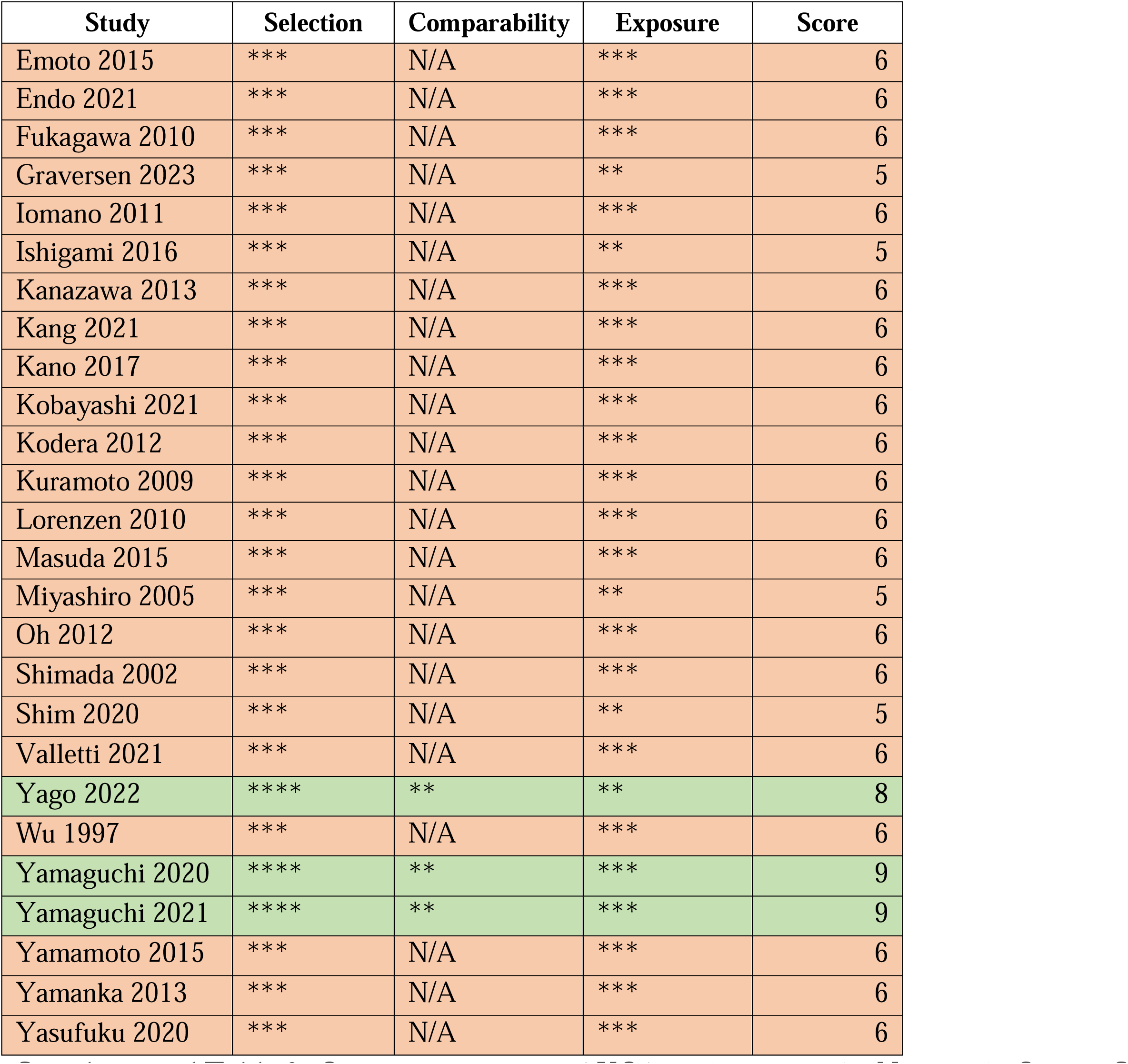
Quality assessment of KQ1 studies using the Newcastle-Ottawa Scale.

| Study | Selection | Comparability | Exposure | Score |
| --- | --- | --- | --- | --- |
| Emoto 2015 | *** | N/A | *** | 6 |
| Endo 2021 | *** | N/A | *** | 6 |
| Fukagawa 2010 | *** | N/A | *** | 6 |
| Graversen 2023 | *** | N/A | ** | 5 |
| Iomano 2011 | *** | N/A | *** | 6 |
| Ishigami 2016 | *** | N/A | ** | 5 |
| Kanazawa 2013 | *** | N/A | *** | 6 |
| Kang 2021 | *** | N/A | *** | 6 |
| Kano 2017 | *** | N/A | *** | 6 |
| Kobayashi 2021 | *** | N/A | *** | 6 |
| Kodera 2012 | *** | N/A | *** | 6 |
| Kuramoto 2009 | *** | N/A | *** | 6 |
| Lorenzen 2010 | *** | N/A | *** | 6 |
| Masuda 2015 | *** | N/A | *** | 6 |
| Miyashiro 2005 | *** | N/A | ** | 5 |
| Oh 2012 | *** | N/A | *** | 6 |
| Shimada 2002 | *** | N/A | *** | 6 |
| Shim 2020 | *** | N/A | ** | 5 |
| Valletti 2021 | *** | N/A | *** | 6 |
| Yago 2022 | **** | ** | ** | 8 |
| Wu 1997 | *** | N/A | *** | 6 |
| Yamaguchi 2020 | **** | ** | *** | 9 |
| Yamaguchi 2021 | **** | ** | *** | 9 |
| Yamamoto 2015 | *** | N/A | *** | 6 |
| Yamanka 2013 | *** | N/A | *** | 6 |
| Yasufuku 2020 | *** | N/A | *** | 6 |

**Supplemental Table 10:**
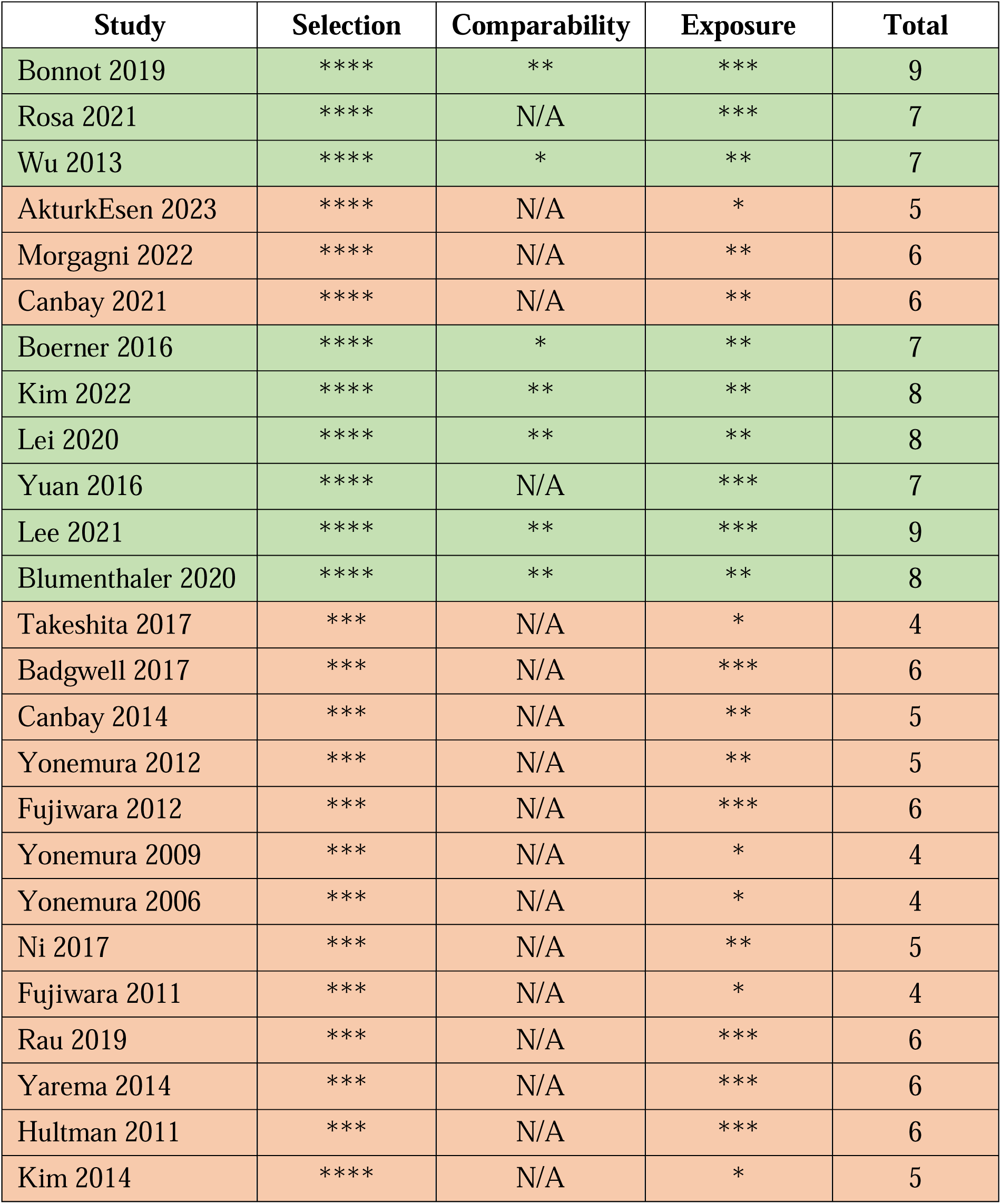
Quality assessment of KQ2 studies using the Newcastle-Ottawa Scale.

**Supplemental Figure 1:**
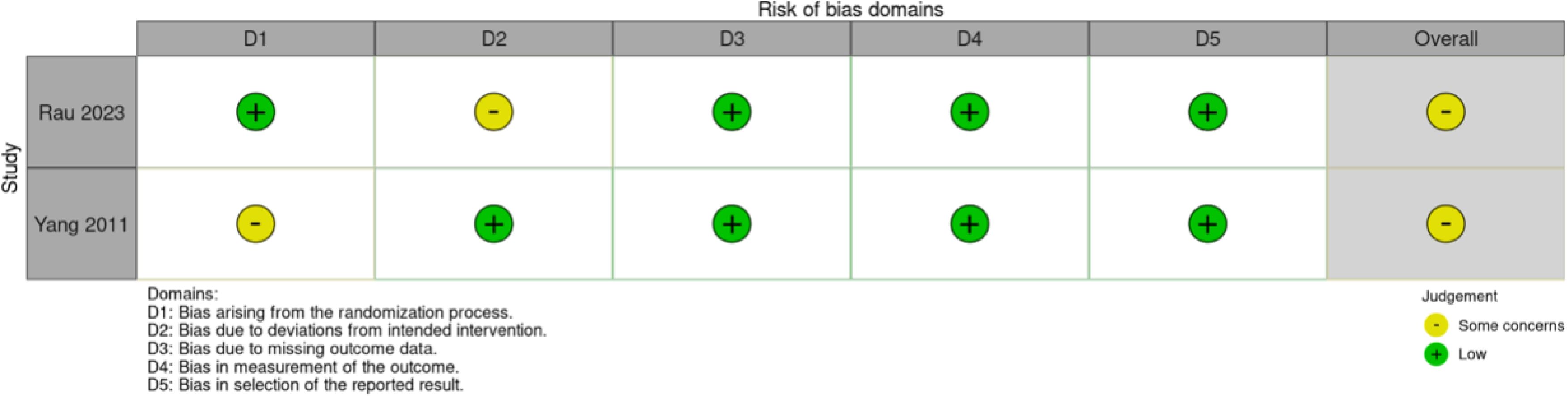
ROB summary figure for two randomized controlled trials evaluating CRS-HIPEC vs. CRS-alone

